# Clinical course and risk factors for in-hospital mortality of 205 patients with SARS-CoV-2 pneumonia in Como, Lombardy Region, Italy

**DOI:** 10.1101/2021.02.25.20134866

**Authors:** Mauro Turrini, Angelo Gardellini, Livia Beretta, Lucia Buzzi, Stefano Ferrario, Sabrina Vasile, Raffaella Clerici, Andrea Colzani, Luigi Liparulo, Giovanni Scognamiglio, Gianni Imperiali, Giovanni Corrado, Antonello Strada, Marco Galletti, Nunzio Castiglione, Claudio Zanon

## Abstract

**Importance:** With randomized clinical trials ongoing and vaccine still a long distance away, efforts to repurpose old medications used for other diseases provide hope for the treatment of COVID-19

**Objectives:** To examine the risk factors for in-hospital mortality and describe the effectiveness of different treatment strategies in a real-life setting of patients with severe acute respiratory syndrome coronavirus 2 (SARS-CoV-2) pneumonia.

**Design:** Real-life single-center study during the Lombardy COVID-19 outbreak.

**Setting:** Valduce Hospital in Como, Lombardy Region, Italy.

**Participants:** 205 laboratory-confirmed patients presenting with SARS-Cov-2 pneumonia requiring hospitalization.

**Interventions:** All patients received best supportive care and, based on their clinical needs and comorbidities, specific interventions that included the main drugs being tested for repurposing to treat COVID-19, such as hydroxychloroquine, anticoagulation, antiviral drugs, steroids or interleukin-6 pathway inhibitors.

**Main outcomes and measures:** Clinical, laboratory and treatment characteristics were analyzed with univariate and multivariate logistic regression methods to explore their impact on in-hospital mortality and compared with current literature data.

**Results:** Univariate analyses for clinical variables showed prognostic significance for age equal or greater than 70 years (estimated 28-days survival: 21.4 vs 67.4%; p<0.0001), presence of 2 or more relevant comorbidities (35.3 vs 61.8%; p=0.0008), ratio of arterial oxygen partial pressure to fractional inspired oxygen (P/F) less than 200 at presentation (21-days survival: 14.7 vs 52.4%;p<0.0001), high levels of lactate dehydrogenase (LDH) (26.4 vs 65.3%; p=0.0001), and elevated C-reactive protein (CRP) values (25.4 vs 74.9%; p=0.0001), while no statistical significance was found for all the other clinical variables tested. At univariate analysis for the different treatment scheduled, prognostic significance for survival was showed for intermediate or therapeutic-dose anticoagulation (estimated 28-days survival: 37.1 vs 23.4%; p=0.0001), hydroxychloroquine (35.7 vs 27.3%; p=0.0029), early antiviral therapy with lopinavir/ritonavir (60.1 vs 22.4%; p<0.0001), late short-course of steroids (47.9 vs 18.2%; p<0.0001) or tocilizumab therapy (69.4 vs 29.4%; p=0.0059). Multivariable regression confirmed increasing odds of in-hospital death associated with age older than 70 years (odds ratio 3.26, 95% CI 1.81–5.86; p<0.0001) and showed a reduction in mortality for patients treated with anticoagulant (−0.37, 0.49-0.95; p=0.0273), antiviral (−1.22, 0.16-0.54; p<0.0001), or steroids (−0.59, 0.35-0.87; p=0.0117) therapy.

**Conclusions and Relevance:** Results from this real-life single-center experience are in agreement and confirm actual literature data on SARS-CoV-2 pneumonia, both in terms of clinical risk factors for in-hospital mortality and as regards the effectiveness of the different therapies proposed for the management of COVID-19 disease. Waiting the results from randomized clinical trials, these data could help clinicians to identify patients with poor prognosis at an early stage and guide the choice between the different treatments implied in COVID-19 disease.

**KEY POINTS:** *Question:* Among the main drugs that have been tested for repurposing to treat COVID-19, what are the most effective medical treatments for SARS-CoV-2 pneumonia?

*Findings:* Results from these real-life cohort of 205 patients confirm at multivariate regression model an increasing odds of in-hospital death associated with age older than 70 years (OR 3.26) and a reduction in mortality for patients treated with anticoagulant (OR −0.37), antiviral lopinavir/ritonavir (OR −1.22), or steroids therapy (OR −0.59). In contrast, hydroxychloroquine and tocilizumab have not been confirmed to have a significant effect in the treatment of SARS-CoV-2 pneumonia, in accordance with the latest data from the international literature.

*Meaning:* Waiting the results from randomized clinical trials, these data could help clinicians to identify patients with poor prognosis at an early stage and guide the choice between the different treatments implied in COVID-19 disease.

## INTRODUCTION

Severe acute respiratory syndrome coronavirus 2 (SARS-CoV-2) causing coronavirus disease 2019 (COVID-19) represents a viral disease infecting more than six million individuals all over the world and has emerged as one of the major public health emergencies of international concern. Therefore, on March 11th 2020 the World Health Organization (WHO) declared COVID-19 a pandemic disease because of widespread infectivity and high contagion rate. Full-genome sequencing indicated that COVID-19 is a betacoronavirus in the same subgenus as the severe acute respiratory syndrome (SARS) virus. The structure of the receptor-binding gene region is very similar to that of the SARS coronavirus, and the virus uses the same receptor, the angiotensin-converting enzyme 2 (ACE2), for cell entry [^1^]. Direct person-to-person transmission is the primary means of transmission of SARS-CoV-2, mainly through close-range contact via respiratory droplets or by transfer to mucous membranes after coming into contact with contaminated surfaces. The median incubation period for COVID-19 is between 2 and 12 days, with most cases occurring approximately four to five days after exposure [^2,3^]. Several studies describing the clinical features of COVID-19 have been performed in hospitalized populations [^4,5,6^]. The most common clinical characteristics at the onset of the disease were fever (even low-grade fever <38° C), cough, fatigue, and dyspnea with typical bilateral infiltrates on chest imaging. Other features, such as upper respiratory tract symptoms, myalgias, diarrhea, abdominal pain, anosmia and dysgeusia, may also be present [^7,8,9,10^]. The spectrum of symptomatic infection ranges from mild to critical, with a proportion of severe or critical disease reported in approximately 20% of cases [^5,11,12^]. Acute respiratory distress syndrome (ARDS) is the major complication in patients with severe disease and can manifest shortly after the onset of dyspnea [^4,21^]. Other common complications include thromboembolic disorders, cardiovascular disease (eg, arrhythmias, acute cardiac injury, and shock), exuberant inflammatory response similar to cytokine release syndrome, and secondary infections [^4,13,14,15,16,17,18^]. Severe illness can occur in otherwise healthy individuals of any age, but the highest proportion of severe cases occurs in adults older than 60 years of age or presenting underlying medical comorbidities. Potential risk factors for severe illness include cardiovascular and cerebrovascular diseases, diabetes mellitus, chronic lung disease, malignancies, and obesity [^19,20,21,22^]. The most common laboratory abnormalities among hospitalized patients with COVID-19 include lymphopenia, elevated aminotransaminase, lactate dehydrogenase (LDH), and inflammatory markers such as C-reactive protein (CRP), ferritin, and erythrocyte sedimentation rate (ESR) [^3,4,6,23,24,25^]. Common abnormal chest radiograph findings were consolidation and ground glass opacities, with bilateral, peripheral, and lower lung zone distributions [^26^]. Chest computed tomography (CT) studies confirm the presence of bilateral peripheral ground-glass opacification with or without consolidative abnormalities, while less common findings include pleural thickening, pleural effusion, and lymphadenopathy [^27,28,29^].

A number of approaches to COVID-19 therapy have been investigated, including chloroquine, antiviral drugs, interleukin-6 pathway inhibitors and other immunomodulators. In vitro studies have shown that hydroxychloroquine and chloroquine can bind cell surface sialic acid and gangliosides with high affinity, thereby impairing SARS-CoV-2 binding to host cell angiotensin converting enzyme (ACE)-2 receptors [^30,31^]. On the basis of their potential antiviral activity in vitro, several randomized trials have been conducted to evaluate their clinical use, but none have suggested a clear efficacy [^32,33,34^]. Most observational studies have also not suggested a benefit with chloroquine or hydroxychloroquine treatment [^35,36^], and highlighted the potential risk for toxicity of those drugs. Arrhythmias and QTc prolongation emerged as the most relevant adverse events, particularly when these agents are administered in patients with comorbidities or in combination with other medicines known to prolong the QT interval, including azithromycin. Antiviral agent lopinavir/ritonavir has been considered as a promising treatment option for COVID-19 infections, based on its proven in vitro efficacy against other novel coronaviruses SARS-COV via inhibition of 3-chymotrypsin-like protease [^37,38^]. However, results from a randomized trial do not demonstrate a clear benefit of lopinavir-ritonavir compared with standard care alone, at the cost of increased toxicity [^39^]. Due to the observation that some patients have a marked elevation in pro-inflammatory markers with a clinical presentation that resembles cytokine release syndrome, interrupting the inflammatory cascade has been proposed as a potential therapeutic target for COVID-19 to prevent disease progression. Tocilizumab, an interleukin(IL)-6 receptor inhibitor, is being evaluated in randomized trials for treatment of COVID-19. Similarly, glucocorticoids have been used in patients with moderate to severe ARDS.

Here, we report the epidemiological, clinical, and laboratory characteristics, treatment and clinical outcomes of 205 laboratory-confirmed cases infected with SARS-CoV-2 admitted to Valduce Hospital in Como, Italy. The aim of this study is to explore risk factors for in-hospital mortality and describe the effectiveness of different treatment strategies in a real-life single-center cohort.

## MATERIALS AND METHODS

### Patients’ characteristics, data collection and treatment protocols

Two hundred and five patients aged between 17 and 100 years (male/female: 113/92) were admitted to Valduce hospital because of SARS-Cov-2 pneumonia requiring hospitalization and were included in this analysis. One hundred and sixty-eight patients came to direct observation in the emergency room, while 37 patients were sent to the hospital from a nursing home for the elderly. All patients had a laboratory-confirmed SARS-CoV-2 infection through RT-PCR on nasopharyngeal swab specimen. For each patient, data regarding history, vital signs, ratio of arterial oxygen partial pressure to fractional inspired oxygen (P/F), need for oxygen therapy, blood chemistry parameters, treatment schedule and outcome were recorded. Blood chemistry variables were recorded both as value at presentation and as maximum value achieved during hospitalization, while interleukin 6 (IL6) values were recorded only in 29 patients candidate for tocilizumab therapy. Patients’ characteristics at presentation are shown in Table 1.

**Table 1.** Clinical characteristics at presentation of patients with SARS-CoV-2 disease

|  | All patients |  | Male |  | Female |  |
| --- | --- | --- | --- | --- | --- | --- |
| Patients, no. | 205 |  | 113 |  | 92 |  |
| Median age, y (IQR) | 77 | (65-83) | 76 | (62-82) | 78 | (70-85) |
| Median P/F ratio (IQR) | 242 | (171-295) | 240 | (171-286) | 248 | (171-309) |
| Median WBC, x 10<sup>9</sup>/L (IQR) | 7.2 | (5.0-9.9) | 7.2 | (5.3-10.1) | 6.7 | (4.7-9.3) |
| Median LDH, U/L (IQR) | 339 | (252-422) | 364 | (276-474) | 294 | (242-381) |
| Median CRP, mg/L (IQR) | 98 | (35-156) | 100 | (45-165) | 82 | (28-137) |
| Median PCT, µg/L (IQR) | 0.15 | (0.07-0.36) | 0.16 | (0.08-0.38) | 0.14 | (0.05-0.28) |
| Median ALT, U/L (IQR) | 27 | (18-46) | 33 | (21-53) | 21 | (17-37) |
| Median AST, U/L (IQR) | 39 | (29-58) | 43 | (31-67) | 36 | (25-49) |

ALT: alanine aminotransferase (normal value: <50); AST: aspartate aminotransferase (normal value: <50); CRP: C-reactive protein (normal value: <5); IQR: interquartile range; LDH: lactate dehydrogenase (normal value: <248); PCT: procalcitonin (normal value: <0,10); P/F ratio: ratio of arterial oxygen partial pressure to fractional inspired oxygen; WBC: white blood cells (normal value: 4,0-10,8)

Median age at presentation was 77 years (interquartile range, IQR: 65-83), with 65 patients (31.7%) aged less than 70 years, 50 patients (24.4%) aged 70 to 79 years, 70 patients (34.1%) aged 80 to 89 years and 20 patients (9.8%) aged 90 years or older. Thirty-five patients (17%) presented no comorbidity, 52 patients (25.4%) had 1 comorbidity, and 118 patients (57.6%) had two or more comorbidities. Among the main comorbidities there were arterial hypertension (n 115), atrial fibrillation (n 43), diabetes (n 32), chronic obstructive pulmonary disease (n 25), and obesity (n 18).

Overall, hydroxychloroquine (HCQ) was administered to 160 patients (78%) with a loading dose of 400 mg bid followed by maintenance with 200 mg die for 7-14 days. Antiviral therapy was administered to 52 patients (25.3%) and consisted in lopinavir/ritonavir 400/100 mg bid for 5-7 days. Since only 2 patients were treated with remdesivir on a compassionate-use basis, this treatment was not included in the statistical analysis. Thirty-four patients (16.5%) continued their usual anticoagulant therapy for previous medical conditions, while 124 patients (60.5%) were treated with low molecular weight heparin (LMWH) at intermediate doses of 100 U/kg/day. Twenty-nine patients were evaluated for anti-IL6 therapy based on their clinical characteristics and drug availability, and 21 patients (10.2%) received tocilizumab 8 mg/kg (up to a maximum of 800 mg per dose) with a second administration after 12 hours on a compassionate use. Ninety patients (43.9%) with the need for oxygen therapy after more than 7 days from the onset of symptoms were treated with steroid therapy, consisting in methylprednisolone 1 mg/kg for 5 days then tapered according to clinical evolution. Almost all patients (95.6%) were treated with a concomitant short course of antibiotic therapy, usually including azithromycin 500 mg die for 3 to 6 days.

The need for oxygen therapy was distinguished based on the amount of oxygen delivered to maintain an adequate P/F ratio. Oxygen delivery systems were classified as low-flow or variable-performance devices and high-flow or fixed-performance devices. Low-flow systems provide oxygen at flow rates that are lower than patients’ inspiratory demands, where high-flow systems provide a constant FiO_2_ by delivering the gas at flow rates that exceed the patient’s peak inspiratory flow. Oxygen was delivered using nasal cannulas (n 32), bag-valve masks (n 36), Venturi mask (n 41) or continuous positive airway pressure (CPAP, n 64). Sixteen patients (7.8%) did not need for oxygen therapy, while 16 patients (7.8%) were transferred to the Intensive Care Unit for orotracheal intubation (IOT).

### Statistical analyses

All collected variables were submitted to descriptive methods. For continuous variables the distribution was firstly evaluated by the Shapiro-Wilk test, so that normally distributed variables were summarized with mean and standard deviation, while non-normal variables were summarized as median, range and interquartile range. Pearson’s chi-square test with Yates’ correction for continuity and Fisher’s exact test (if applicable) were used to check the association between categorical data, after cross-tabulation. Comparisons of normally distributed continuous variables were carried out by Student’s t-test or by Welch’s test (in the case of non-homogeneous variances between groups, previously verified by Levene’s test). The Mann-Whitney U test was used for comparison of continuous non-normally distributed variables. The survival analysis was carried out using the Kaplan-Meier product limit method, followed by the logrank test, to evaluate the possible differences in survival between groups. Cox univariate and multivariate regression models were also used to analyse the effects of continuous variables on survivorship. The optimal multivariate model was chosen using a backward stepwise elimination after inserting all variables showing *p* < 0.05 at univariate analysis. The receiver operating characteristics curve (ROC) was traced to analyse the role of continuous variables on survivorship and to search for an optimal cut-off value for variables themself. For all possible cut-off points, the total accuracy was considered together with sensitivity, specificity, positive predictive value and negative predictive value; however, the choice was made according to Youden. Statistical significance was assumed for all tests with *p* <.05. Statistical analysis was done using MedCal 9.3.7.0.

## RESULTS

Two hundred and five patients were included in the analysis. The median follow-up time was 16 days based on the reverse Kaplan-Meier method. The median hospitalization time was 7 days (range: 1-27) and 12 days (range: 1-43) in deceased and surviving patients, respectively. Among the 107 surviving patients (52.2%), 58 were discharged from the hospital and 49 were transferred to rehabilitation facilities. The estimated 7-days and 28-days survival resulted 69.4% and 34.1%, respectively (Figure 1). Statistical analysis showed no significant difference in term of clinical variables between male and female patients.

**Figure 1.**
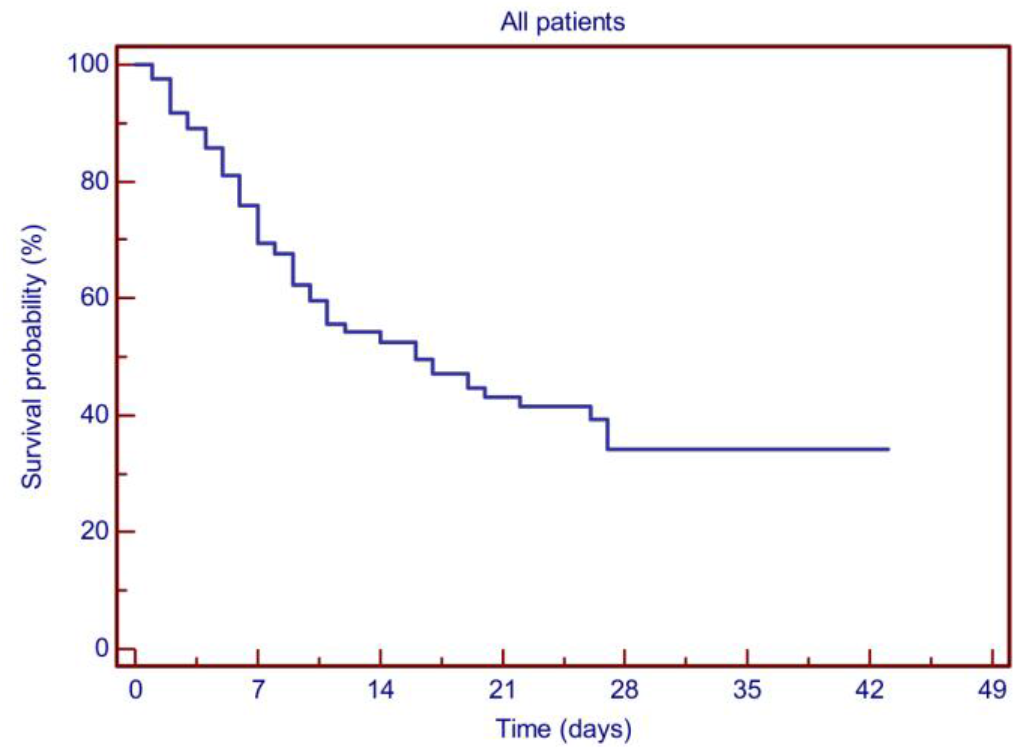
Kaplan-Meier plots showing the probability of survival for the whole patient population.

### Prognostic factors for survival

Cox univariate and multivariate regression models were performed for all patients to evaluate the role of different clinical variables as predictors for in-hospital mortality. The following potential prognostic parameters were evaluated: age, sex, comorbidities, P/F ratio, oxygen therapy, white blood cell count (WBC), lactate dehydrogenase (LDH), C-reactive protein (CRP), procalcitonin (PCT), serum ferritin, D-dimer, interleukin 6 (IL6), alanine aminotransferase (ALT), and aspartate aminotransferase (AST). Blood chemistry variables were assessed for survival both as value at presentation and as maximum value achieved during hospitalization.

For continuous variables, a ROC curve analysis was performed towards survival in search of possible cut-off values. Age distribution and P/F ratio at presentation showed an optimal cut-point at 69 years (AUC 0.71, sensitivity 86.7%, specificity 48.6%, LR+ 1.69, LR-0.27; p=0.0001) and at 233 (AUC 0.73, sensitivity 65.4%, specificity 75.0%, LR+ 2.62, LR-0.46; p=0.0001), respectively. Among the surrogate markers of inflammation, LDH and CRP showed an optimal cut-point at 395 U/L (AUC 0.77, sensitivity 75.0%, specificity 71.2%, LR+ 2.61, LR-0.35; p=0.0001) and at 124 mg/L (AUC 0.70, sensitivity 80.3%, specificity 60.9%, LR+ 2.05, LR-0.32; p=0.0001) respectively, while no significant cut-off points for WBC, PCT, ferritin, D-dimer, ALT or AST were identified. Between the 29 patients tested, IL6 values showed an optimal cut-point at 3484 pg/mL (AUC 0.862, sensitivity 66.7%, specificity 95.6%, LR+ 15.33, LR-0.35; p=0.0003).

At univariate analyses, age with cut-off point set at 70 years showed prognostic significance for survival, with an estimated 28-days survival of 67.4% and 21.4% for patients aged less or greater than 70 years, respectively (logrank test: p<0.0001)(Figure 2A). Univariate analyses for clinical variables showed prognostic significance for the number of relevant comorbidities (28-days survival: 61.8%, 51.7% and 35.3% for none, 1 or 2 or more comorbidities; p=0.0008), P/F ratio less than 200 at presentation (21-days survival: 14.7% vs 52.4%; p<0.0001), high levels of LDH (28-days survival: 26.4% vs 65.3%; p=0.0001), and elevated C-reactive protein (CRP) values (25.4% vs 74.9%; p=0.0001), while no statistical significance was found for all the other clinical variables tested (see Figure 3A-D).

**Figure 2.**
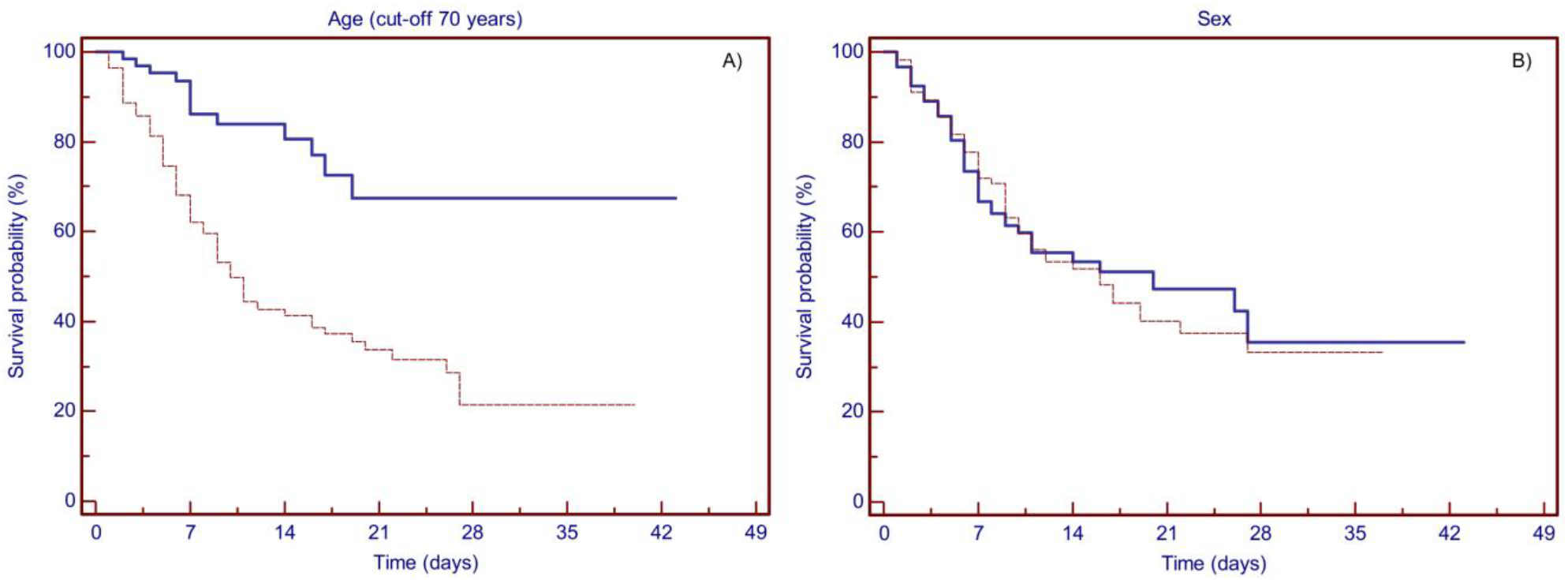
Kaplan-Meier plots showing the probability of survival based on patients’ characteristics. (A) Patients aged less than 70 years (solid line) or older than or equal to 70 years (dashed line). Age showed prognostic significance for survival at a cut-off point set at 70 years (p<0.0001), with an estimated 28-days survival of 67.4% and 21.4%, respectively. (B) Survival was not affected by the gender (female, solid line; male, dashed line)(p=0.086).

**Figure 3.**
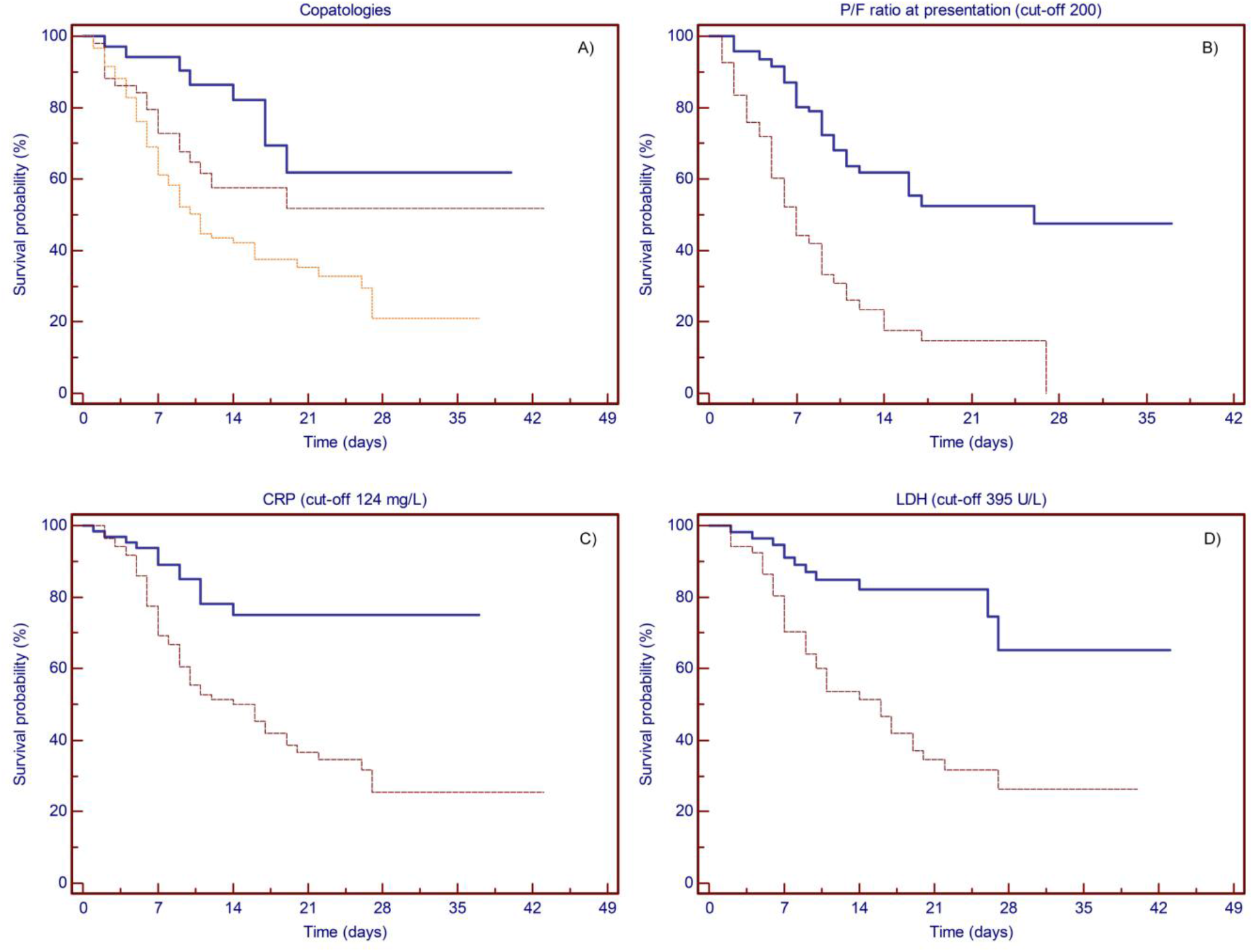
Kaplan-Meier plots showing the probability of survival based on clinical variables. (A) Patients with no comorbidity (solid line), 1 comorbidity (dashed line), and 2 or more comorbidities (dotted line). The number of comorbidities showed prognostic significance for survival (p=0.0008), with an estimated 28-days survival of 61.8%, and 51.7% and 35.3%, respectively. (B) P/F ratio at presentation more than 200 (solid line) or less than or equal to 200 (dashed line). P/F ratio at presentation showed prognostic significance for survival at a cut-off point set at 200 (p<0.0001), with an estimated 21-days survival of 52.4% and 14.7%, respectively. (C) CRP less than 124 (solid line) or more than or equal to 124 (dashed line). CRP values showed prognostic significance for survival at a cut-off point set at 124 mg/L (p=0.0001), with an estimated 28-days survival of 74.9% and 25.4%, respectively. (D) LDH less than 395 (solid line) or more than or equal to 395 (dashed line). LDH values showed prognostic significance for survival at a cut-off point set at 395 U/L (p=0.0001), with an estimated 28-days survival of 65.3% and 26.4%, respectively.

Finally, univariate analysis was tested for survival for the different treatment scheduled (Figure 4A-F). Patients requiring only low-flow oxygen therapy experienced improved survival compared to patients deserving of high-flow therapy, with an estimated 28-days survival of 50.0% and 23.3%, respectively (p<0.0001). The use of any anticoagulant treatment resulted in an improvement in survival (estimated 28-days survival: 37.5 vs 23.8%; p=0.0001), while no difference was recorded between intermediate-dose or therapeutic-doses anticoagulation (36.9% and 37.1%, respectively). Hydroxychloroquine treatment was administered to 160 patients, resulting in an estimated 28-days survival of 35.7% vs 27.3% of those who did not receive such therapy (*p=*0.0029). Fifty-two patients received early antiviral therapy with lopinavir/ritonavir, which resulted in an estimated 28-days survival of 60.1% vs 22.4% (*p*<0.0001). Ninety patients were treated with steroid and 21 patients received tocilizumab. Both anti-inflammatory treatments showed prognostic significance for survival, with an estimated 28-days survival between treated and not-treated patients of 47.9% vs 18.2% (*p*<0.0001) and 69.4% vs 29,6% (*p*<0.0059), respectively.

**Figure 4.**
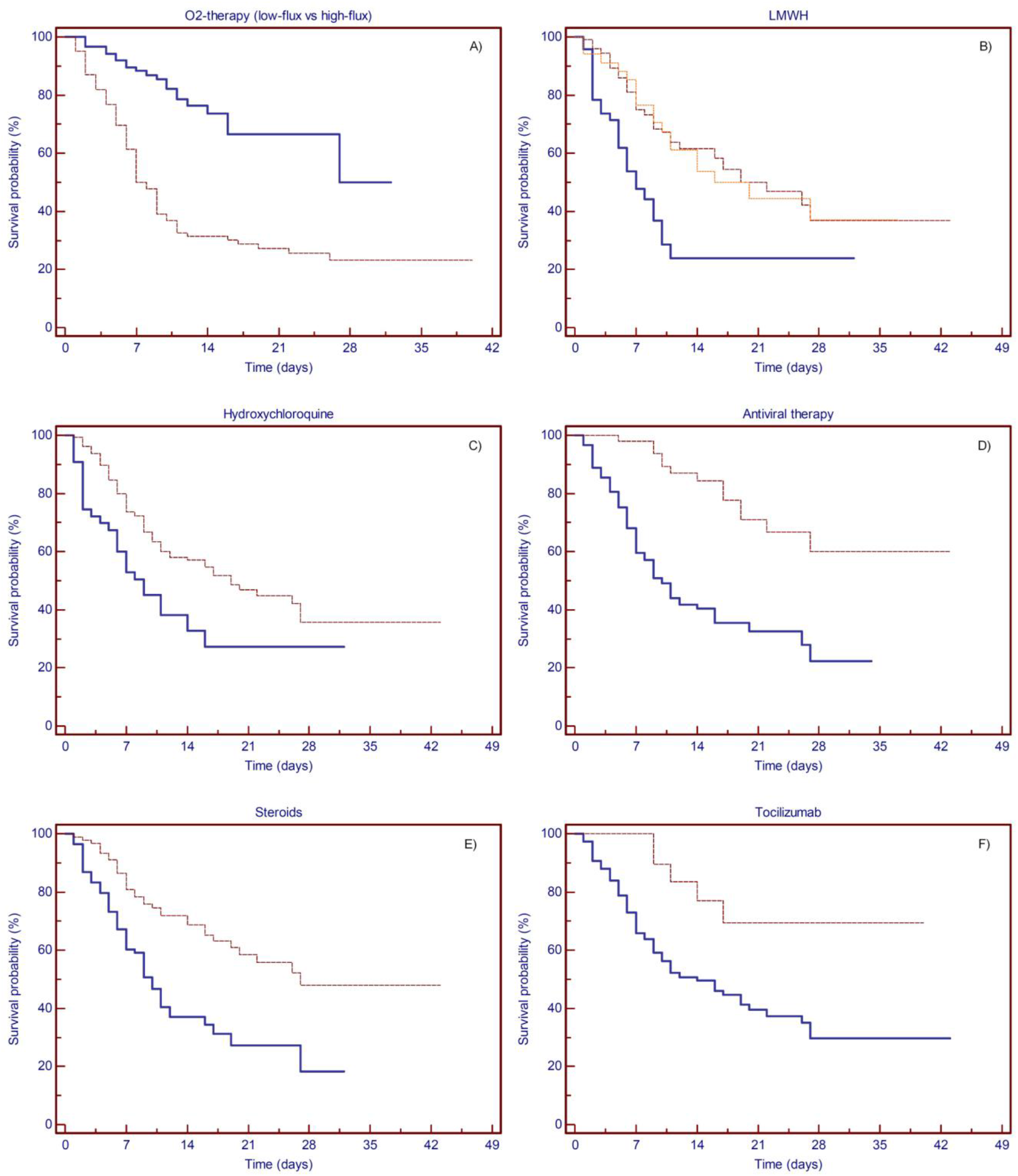
Kaplan-Meier plots showing the probability of survival based on treatments. (A) Patients treated with low-flux (solid line) or high-flux (dashed line) oxygen-therapy. O2-therapy showed prognostic significance for survival (p<0.0001), with an estimated 28-days survival of 50.0%, and 23.3%, respectively. (B) No anticoagulation therapy (solid line), LMWH at prophylaxis dose (dashed line) or therapeutic dose (dotted line). LMWH treatment showed prognostic significance for survival (p=0.0001), with an estimated 28-days survival of 23.8%, 36.9% and 37.1%, respectively. (C) No HCQ (solid line) vs HCQ treatment (dashed line). HCQ showed prognostic significance for survival (p=0.0029), with an estimated 28-days survival of 27.3% and 35.7%, respectively. (D) No antiviral (solid line) vs antiviral treatment (dashed line). Antiviral therapy showed prognostic significance for survival (p<0.0001), with an estimated 28-days survival of 22.4% and 60.1%, respectively. (E) No steroid (solid line) vs steroid treatment (dashed line). Steroids therapy showed prognostic significance for survival (p<0.0001), with an estimated 28-days survival of 18.2% and 47.9%, respectively. (F) No tocilizumab (solid line) vs tocilizumab treatment (dashed line). Tocilizumab therapy showed prognostic significance for survival (p =0.0059), with an estimated 28-days survival of 29.6% and 69.4%, respectively.

When combined in the multivariate analysis with backward elimination of factors, multivariable regression confirmed increasing odds of in-hospital death associated with age older than 70 years (OR 3.26, 95% CI 1.81–5.86; p<0.0001) and showed a reduced odds of mortality in patients treated with intermediate or therapeutic dose anticoagulation (OR −0.37, 95% CI 0.49-0.95; p=0.0273), antiviral drug lopinavir/ritonavir (OR −1.22, 95% CI 0.16-0.54; p<0.0001), or steroids therapy (OR −0.59, 95% CI 0.35-0.87; p=0.0117) (Cox proportional-hazards regression model: *p*<0.0001) (Table 2).

**Table 2.** Multivariate analysis.

| Variable | b | SE | Exp(b) | 95% CI of Exp(b) | p |
| --- | --- | --- | --- | --- | --- |
| Age (cut-off 70y) | 1,1821 | 0,3011 | 3,2612 | 1,8129 to 5,8665 | < 0,0001 |
| Anticoagulation | -0,3736 | 0,1692 | 0,6883 | 0,4948 to 0,9574 | 0,0273 |
| Antiviral therapy | -1,2202 | 0,3106 | 0,2952 | 0,1611 to 0,5409 | < 0,0001 |
| Steroid therapy | -0,5919 | 0,2347 | 0,5533 | 0,3501 to 0,8744 | 0,0117 |

### Prognostic score model

Based on ROC curve analysis, inflammation markers CRP and LDH were incorporated into a prognostic score system, attributing 1 point for each value above the identified cut-off (nominally, 124 mg/L for CRP and 395 U/L for LDH). Having none, one or both of the positive markers determined a reduction in estimated 28-days survival from 94.0% to 51.3% and 21.0%, respectively (logrank test: p < 0.0001)(Figure 5).

**Figure 5.**
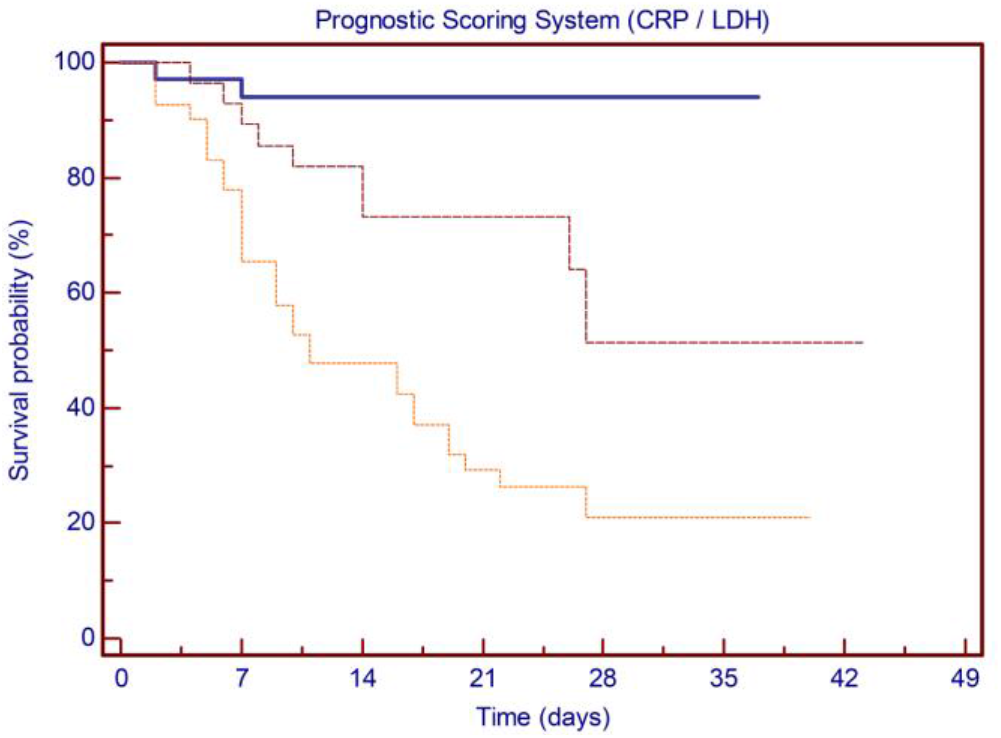
Kaplan-Meier plots showing the probability of survival based on prognostic score model. Patients presenting none (solid line), one (dashed line) or both (dotted line) of the positive markers. Estimated 28-days survival varies from 94.0% to 51.3% and to 21.0%, respectively (p<0.0001).

## DISCUSSION

At the end of 2019, a novel coronavirus was identified as the cause of a cluster of pneumonia cases in the region of Wuhan, China. The infection rapidly spread, resulting in an epidemic throughout China, followed by an increasing number of cases in other countries throughout the world. Individuals of any age can acquire SARS-CoV-2 infection, although middle-aged and older adults are most commonly affected [^4,5^]. In a modeling study based on Chinese data, the hospitalization rate for COVID-19 increased with age, with a 11.8% rate for those 60 to 69 years old, 16.6% rate for those 70 to 79 years old, and 18% for those older than 80 years [^40^]. Older age has also been associated with more severe disease and increased fatality rate, though there is no clear age cut-off [^11,22,41^]. In a report from the Chinese Center for Disease Control and Prevention, mortality rates were 8 and 15% among those aged 70 to 79 years and 80 years or older, respectively, in contrast to the 2.3% case fatality rate among the entire cohort [^11^]. Similar findings were reported from Onder et al. in an Italian cohort, with case fatality rates of 12.8 and 20.2% among patients aged 70 to 79 years and 80 years or older, respectively [^22^].

Among the approximately 4,000 confirmed infections reported to CDC in the United States, mortality was confirmed highest among older individuals, with 80% of deaths occurring in person over the age of 65 and with the highest percentage of severe outcomes among those older than 85 years [^42^]. Our cohort of patients is consistent with this data, with a median age at presentation of 77 years and 68.2% of patients aged 70 or older. Consistent with CDC data, mortality was confirmed highest among elderly patients, with 86.7% and 62.2% of deaths occurring in person older than 70 and 80 years, respectively. Analysis of ROC curve confirmed an optimal cut-point at 69 years, with an estimated 28-day survival of 67.4% and 21.4% for patients younger or older than 70 years, and age over 70 years proved to be an independent risk factors for survival when combined in the multivariate analysis. Several comorbidities have been associated with severe illness and mortality in SARS-CoV-2 infection. In a subset of 355 Italian patients who died with COVID-19, the mean number of preexisting comorbidities was 2.7, with less than 1% of patients presenting no underlying condition and approximately half of patients presenting with 3 or more comorbidities [^22^]. In accordance with these data, in our study 57.6% of patients had two or more preexisting comorbidities among those considered potential risk factors for severe COIVD-19 disease. Survival analysis showed prognostic significance for comorbidities, but the significance was not retained when tested against other clinical and therapeutic variables in multivariate analysis.

Several laboratory features may be altered among hospitalized patients with COVID-19. High D-dimer levels and severe lymphopenia have been associated with critical illness, while procalcitonin levels on admission are more likely to be elevated in patients requiring ICU care [^4,5,7^]. Elevated lactate dehydrogenase, C-reactive protein and interleukin-6 levels are other common laboratory findings among hospitalized patients with COVID-19 [^3,4,6,24^]. However, none of these characteristics have clearly demonstrated to have a strong prognostic value. In our patient population LDH, CRP and IL-6 showed an optimal cut-point for survival on ROC curve analysis, while we found no significance for the other laboratory test WBC, PCT, ferritin, and D-dimer. Based on these data, inflammation markers CRP and LDH have been incorporated into a prognostic score system, which has been shown to recognize 3 distinct groups of patients characterized by different disease severity and clinical outcome. Validation of this potential easy-to-apply clinical score is required on a larger prospective patient population.

The rapid diffusivity and high mortality of severe cases have led researchers and clinicians to experience the impact of new and old drugs for the treatment of SARS-CoV-2 infection. The optimal approach to treatment of COVID-19 disease is uncertain, since for most potential therapies evidence for their use comes primarily from observational data or indirect evidence. Hydroxychloroquine (HCQ), notably used in autoimmune diseases, has been considered of potentially interest in SARS-CoV-2 infection. In vitro studies have indeed shown that HCQ binds cell surface sialic acid and gangliosides with high affinity, thereby impairing SARS-CoV-2 spike protein recognition and binding to host cell ACE-2 receptors [^30^]. On the basis of these data, the Taiwan CDC declared HCQ a potential important anti-SARS-CoV-2 agent on 26 March, 2020. An early French study highlighted the ability of 600 mg daily of HCQ, particularly in combination with azithromycin, in clearing the respiratory viral loads in 3 to 6 days [^32^]. A subsequent retrospective multicenter cohort study among 1438 hospitalized patients with a diagnosis of COVID-19 hospitalized in metropolitan New York, treatment with hydroxychloroquine, azithromycin, or both, compared with neither treatment, was not significantly associated with differences in in-hospital mortality [^33^]. Finally, in an open-label randomized trial of 150 hospitalized patients with mild to moderate COVID-19, adding hydroxychloroquine to standard of care did not improve the rate of SARS-CoV-2 clearance nor resulted in symptomatic improvement by 28 days [^34^]. In accordance with these literature data, in our patient cohort HCQ was safely administered to 160 patients and no treatment-related adverse events were recorded. Although HCQ treatment showed prognostic significance for survival at univariate analysis, significance was not confirmed in the multivariate analysis (*p* 0.146). Finally, several Drug Agencies from different countries now recommend not to use hydroxychloroquine or chloroquine outside the setting of a clinical trial, given the lack of clear benefit from limited data and potential for toxicity. A multi-centre, adaptive, randomized, open clinical trial of the safety and efficacy of treatments for COVID-19, including HCQ, in hospitalized adults is now ongoing (NCT04315948).

The combined protease inhibitor lopinavir/ritonavir, primarily used for HIV infection, has demonstrated an in vitro activity against the SARS-CoV via inhibition of 3-chymotrypsin-like protease [^37,38^]. In a multicentric retrospective matched cohort study conducted during the SARS outbreak the addition of lopinavir/ritonavir as initial treatment was associated with a statistically significant reduction in the overall death rate and intubation rate compared with matched controls [^43^]. In addition, a significant reduction in adverse events such as severe respiratory deterioration and lower nosocomial infections were also noted in patients treated with lopinavir/ritonavir, and significance was confirmed at multivariate analysis [^37,43^]. Notably, no published in vitro activity data against the SARS-CoV-2 exist for Lopinavir/ritonavir. Results from a randomized open-label trial involving hospitalized adult patients with confirmed SARS-CoV-2 infection did not demonstrate a clear benefit of lopinavir/ritonavir treatment in 199 patients with severe COVID-19 beyond standard care alone [^39^]. Though there was a trend towards decreased mortality (particularly when administered within 12 days of symptom onset) and a reduction in serious adverse events, treatment with lopinavir/ritonavir was not associated with a difference in the time to clinical improvement nor in mortality at 28 days. Furthermore, the timing of lopinavir/ritonavir administration during the early peak of viral replication phase (initial 7-10 days) appears to be crucial, since delayed therapy initiation showed no effect on clinical outcomes [^43,44^].

In our study, short-term antiviral therapy was administered to 52 patients early on admission and within 7 days of symptom onset. Furthermore, particular attention had been paid to possible drug-drug interactions, with only a few low-grade transient gastrointestinal events and no serious treatment-related adverse. At univariate analysis, antiviral therapy showed prognostic significance for survival, with a clear benefit in term of 28-days survival. When combined in the multivariate analysis, antiviral therapy proved to be an independent risk factors for survival. The WHO has recently launched the multinational SOLIDARITY trial to further evaluate the activity of those anti-viral treatment in COVID-19 disease.

Markedly elevated inflammatory markers (eg, D-dimer, ferritin) and elevated pro-inflammatory cytokines as interleukin (IL)-6 are associated with severe COVID-19 disease, and blocking the inflammatory pathway has been hypothesized to prevent disease progression [^15^]. Glucocorticoids were the first drugs introduced with this aim [^5,7^]. Based on data reporting potential benefit of glucocorticoids in patients with moderate to severe ARDS, a weak recommendation in favor of such therapy has been provided in this population. However, glucocorticoids administration in critically ill patients with COVID-19-related ARDS is not suggest routinely, since data are limited to a single retrospective Chinese cohort [^12^] and there is concerning about a delayed viral clearance in patients with ARDS due to viral pneumonia. In our cohort, glucocorticoids were administered in patients still needing oxygen therapy at the end of the viral replication phase (ie, after more than 7 days from the onset of symptoms), and consisted in a short course of methylprednisolone up to 1 mg/kg for 5 days with a rapid tapering. Patients who presented with symptoms or signs of bacterial over-infection were excluded from glucocorticoids treatment. At univariate analysis, corticoids therapy showed a clear survival benefit with a rapid improvement of the respiratory pattern, and when combined in the multivariate analysis glucocorticoids treatment proved to be an independent risk factors for survival.

Tocilizumab is an IL-6 receptor inhibitor used for rheumatic diseases and cytokine release syndrome (CRS) in CAR-T therapy. Several case reports and observational studies have described the use of tocilizumab in patients with COVID-19 [^45,46,47,48^], and its benefits have been evaluated in a prospective open, single-arm multicentre study on patients with severe disease [^49^]. No major adverse events have been directly related to tocilizumab treatment, and its use was associated with a decrease in inflammatory markers and improvement in respiratory parameters overall with a reduce number of ICU admissions. A prospective series of 100 consecutive COVID-19 Italian patients with pneumonia and ARDS confirmed that the response to tocilizumab was rapid, sustained, and associated with significant clinical improvement [^50^].

In our patient cohort tocilizumab was administered for compassionate use to 21 patients with moderate to severe COVID-19 pneumonia. No evident treatment-related adverse events were recorded, and although tocilizumab infusion showed a prognostic significance for 28-days survival at univariate analysis, significance was not confirmed in the multivariate analysis (p 0.8716). Notably, we observed that inflammatory markers and IL-6 levels varied heterogeneously after tocilizumab therapy, and patients with increasing levels after treatment did not present a clinical or respiratory worsening. Results from TOCIVID-19 trial, a multicenter, single-arm, open-label, phase 2 study on the efficacy and tolerability of tocilizumab in the treatment of patients with COVID-19 pneumonia (NCT04317092), will soon be available. Furthermore, an open-label, randomized, multicenter study on the efficacy of early compared to late administration of tocilizumab in reducing the number of patients with COVID-19 pneumonia who require mechanical ventilation is now ongoing (NCT04346355).

Abnormalities in coagulation testing have been observed in nearly 20% of patients with COVID-19 disease, and heparin has been recommended by several expert societies because of the risk of venous thromboembolism. Recently, Giardini et al. identified a possible mechanism between SARS-CoV-2 infection and the prothrombotic profile observed in these patients. Data offered a link between ACE2 downregulation and an AngII/s-Flt-1 mediated endothelial disfunction, in a model that strictly resembles preeclampsia and which could offer an explanation to the pathogenesis of acute global vascular damage observed in these patients [^51^]. Several studies suggest a high rate of thromboembolic complications among hospitalized patients, particularly those with presenting with a severe disease [^5,52,53,54^]. In a retrospective series of more than 2500 hospitalized patients with COVID-19, anticoagulation was associated with improved in-hospital survival in intubated patients, with no evident increment of bleeding events [^55^]. In a retrospective study of individuals with severe COVID-19, LMWH appeared to be associated with improved survival when compared with no pharmacologic prophylaxis, especially in patients with high D-dimer levels [^56^]. Reports from ICU patients with severe COVID-19 suggest a higher incidence of venous thrombosis embolism, even using standard prophylaxis [^57,58^]. The question of adequate anticoagulation dose for thromboprophylaxis has also been raised in COVID-19 patients. There are no actual data comparing prospectively the different levels of anticoagulation (prophylactic, intermediate, or therapeutic dosing) in COVID-19 patients, and clinical trials are ongoing. In our study population we favor intermediate-dose anticoagulation (LMWH 100 U/Kg/d) as pharmacologic prophylaxis of venous thromboembolism for hospitalized patients with COVID-19. Interestingly, we found no difference in term of 28-days survival between patients treated with intermediate-dose or therapeutic-dose anticoagulation for previous medical issues. In contrast, thromboprophylaxis at any dose appears to be associated with improved survival when compared with no pharmacologic prophylaxis, and significance is maintained even when combined in multivariate analysis.

Our study has several limitations due to the retrospective nature of the study. Overall, we found that age at the onset of SARS-CoV-2 disease is a powerful predictor of in-hospital mortality. In addition, intermediate or therapeutic-dose anticoagulation, early short-term antiviral therapy and short course of corticosteroids at the end of the viral replication phase proved to be effective treatments in COVID-19 disease. Interestingly, hydroxychloroquine therapy has not been confirmed as significant in the treatment of SARS-CoV-2 infection, in accordance with latest literature data. Finally, impairment in inflammation markers can find application as predictors of poor outcomes for hospitalized patients when incorporated into an easy-to-use prognostic score, helping clinicians to identify patients with poor prognosis at an early stage. Confirmation from a validation cohort is expected. Results from this real-life single-center experience are in agreement and confirm actual literature data on SARS-CoV-2 pneumonia, both in terms of risk factors for in-hospital mortality and as regards the effectiveness of the different therapies proposed for the management of COVID-19 disease. Results from randomized clinical trials are expected.

## Data Availability

The data that have been collected for this study are available from the corresponding author on reasonable request. Participant data without identifiers will be made available after approval from the corresponding author. The proposal with detailed description of study objectives and statistical analysis plan will be needed for evaluation of the reasonability to request for our data. The corresponding author will make a decision based on these materials.

## ACKNOWLEDGMENTS

ZC conceived the study and is responsible for the overall content as guarantor. BL and BL collected the data and provided operational support. TM validated data collection, performed the literature search, conducted the statistical analyses, and drafted the manuscript. GA performed the literature search, contributed in drafting the manuscript and to critical revision of the report. All authors were involved in the clinical management of patients, discussed the results and approved the final manuscript. The corresponding author attests that all listed authors meet authorship criteria.

All authors declare: no support from any organisation for the submitted work; no financial relationships with any organisations that might have an interest in the submitted work in the previous three years; no other relationships or activities that could appear to have influenced the submitted work.

We acknowledge the Valduce Hospital COVID-19 task force and all clinicians, nurses and health-care workers involved in the diagnosis and treatment of patients in Como, Italy.

